# Transmission and clinical characteristics of coronavirus disease 2019 in 104 outside-Wuhan patients, China

**DOI:** 10.1101/2020.03.04.20026005

**Authors:** Chengfeng Qiu, Qian Xiao, Xin Liao, Ziwei Deng, Huiwen Liu, Yuanlu Shu, Dinghui Zhou, Ye Deng, Hongqiang Wang, Xiang Zhao, Jianliang Zhou, Jin Wang, Zhihua Shi, Da Long

**Author notes:** **Corresponding Author:** Chengfeng Qiu, Dr,; Da Long, Prof. Chengfeng Qiu, Qian Xiao, Xin Liao, Ziwei Deng, Huiwen Liu, Yuanlu Shu, Dinghui Zhou, Ye Deng, Hongqiang Wang contributed equally to this article. Address reprint requests to Dr. Qiu at the First People’s Hospital of Huaihua, Huaihua, Hunan, China, or at; to Prof. Da Long at the Shaoyang Central Hospital, Shaoyang, Hunan, China, or at, respectively.

## Abstract

**Background:** Cases with coronavirus disease 2019 (COVID-19) emigrated from Wuhan escalated the risk of spreading in other cities. This report focused on the outside-Wuhan patients to assess the transmission and clinical characteristics of this illness.

**Methods:** Contact investigation was conducted on each patient who admitted to the assigned hospitals in Hunan Province (geographically adjacent to Wuhan) from Jan 22, 2020 to Feb 12, 2020. Demographic, clinical, laboratory and radiological characteristics, medication therapy and outcomes were collected and analyzed. Patients were confirmed by PCR test.

**Results:** Of the 104 patients, 48 (46.15%) were imported cases and 56 (53.85%) were indigenous cases; 93 (89.42%) had a definite contact history with infections. Family clusters were the major body of patients. Transmission along the chain of 3 “generations” was observed. Mean age was 43 (rang, 8-84) years (including 3 children) and 49 (47.12%) were male. Most patients had typical symptoms, 5 asymptomatic infections were found and 2 of them infected their relatives. The median incubation period was 6 (rang, 1-32) days, of 8 patients ranged from 18 to 32 days. Just 9 of 16 severe patients required ICU care. Until Feb 12, 2020, 40 (38.46%) discharged and 1 (0.96%) died. For the antiviral treatment, 80 (76.92%) patients received traditional Chinese medicine therapy.

**Conclusions:** Family but not community transmission occupied the main body of infections in the two centers. Asymptomatic transmission demonstrated here warned us that it may bring more risk to the spread of COVID-19. The incubation period of 8 patients exceeded 14 days.

## Introduction

Coronavirus disease 2019 (COVID-19) was officially named by the World Health Organization (WHO) on Feb 11, 2020. As the causative pathogen of novel coronavirus pneumonia (NCP), COVID-19 has caused more than 67081 patients worldwide^1^. Evidences have demonstrated the person-to-person transmission of COVID-19^2-4^. The controversy of the sharply increased infections and the medical shortage in Wuhan may be the most reason leading to the outbreak of COVID-19 in early stage. On Jan 23, 2019 before the coming of Chinese Lunar New Year, a precedent scale of Wuhan Shutdown was implemented to block or slow the spread of COVID-19.

Unlike the initial infections which all closely related to the Huanan seafood market in Wuhan, China, infections in other cities mainly linked to the patients emigration from Wuhan^5^. It’s worth noting that the epidemic and clinical status of outside-Wuhan may largely differ with the observed status in Wuhan. Serval studies have showed the high ICU rates, hospital-associated infections in Wuhan^2,6^. Until we know this information about COVID-19, it also hard to assess how bad this novel coronavirus is going to get.

Hunan Province geographically adjacent to Wuhan, Hubei Province, high-efficiency transport between the two provinces may lead to a rapid spread of COVID-19 in Hunan Province. This report included the hospitalized patients with COVID-19 to assess the transmission and clinical characteristic of two hospitals, which designated as the treatment center for the NCP in Huaihua and Shaoyang cities, Hunan Province, China. These findings could provide value information to better understand such new illness.

## Methods

### Study population

In this study, we recruited confirmed patients with COVID-19 from two hospitals, the First People’s Hospital of Huaihua and the Central Hospital of Shaoyang which designated as the treatment center of Huaihua and Shaoyang city, Huanan Province, China from Jan 22, 2020 to Feb 12, 2020. According to the guidelines of China^7^, patient was confirmed by the positive result from the real-time reverse-transcription-polymerase-chain-reaction (RT-PCR) assay of nasopharyngeal or throat swab. Suspected infectors that did not confirmed by PCR were excluded. For the study population, imported case was defined as an infector who emigrated from Wuhan (who ever lived in or traveled to Wuhan), the rest of study patients were defined as indigenous cases.

### Procedures

We carefully surveyed the contact history of every patients, including whether he or she ever lived in or travelled to Wuhan, or had closely contacted with people returning from Wuhan during two months before their illness onset. In addition, the history of contacting with animals and eating game meat was also screened. If necessary, we directly communicated with the attending physician, patients or their family members. Demographic, clinical, laboratory and radiological characteristics, medication therapy (ie, antiviral therapy, antibacterial therapy, corticosteroid therapy and traditional Chinese medicine therapy), underlying comorbidities, symptoms, sign and chest computed tomographic images were obtained from electronic medical records. Outcomes were followed till Feb 12, 2020. Standard questionnaire and form were used for contact investigation and data collection. The data were independently reviewed by two trained physicians (Ye Deng and Xin Liao) and checked by another two physicians (Hongqiang Wang and Da Long) respectively. Every one signed Data Authenticity Commitment and stamp official seal.

The date of onset symptom was defined as the day when the case firstly developed symptoms related to NCP. Acute respiratory distress syndrome (ARDS) was defined according to the Berlin definition^8^. Acute kidney injury was identified by an abrupt decrease in kidney function including changes in serum creatinine (SCr) (≥0.3mg/dl or 265.5μmol/L) when they occur within a 48-hour period, other diagnostic items according to evaluation, and management of acute kidney injury: a KDIGO summary^9^. Liver function abnormal was defined as abnormal of liver enzymes or bilirubin. Cardiac injury was identified by the serum levels of cardiac biomarkers (eg, troponin I) which is above the 99^th^ percentile upper reference limit or new abnormalities shown in electrocardiography and echocardiography^2^.

### Real-Time reverse transcription polymerase chain reaction assay

In this study, case confirmation accords to the positive results of PCR. Nasopharyngeal swab was collected from suspected patients. Sample collection and extraction followed the standard procedure. The primers and probe target to open reading frame (ORF1ab) and nucleoprotein (N) gene of COVID-19 were used.

Target 1(ORF1ab):

Forward primer: CCCTGTGGGTTTTACACTTAA,

Reverse primer: ACGATTGTGCATCAGCTGA,

The probe: 5’-FAM-CCGTCTGCGGTATGTGGAAAGGTTATGG-BHQ1-3’;

Target 2 (N):

Forward primer: GGGGAACTTCTCCTGCTAGAAT,

Reverse primer: CAGACATTTTGCTCTCAAGCTG,

The probe: 5’-FAM-TTGCTGCTGCTTGACAGATT-TAMRA-3.

The procedure and reaction condition for PCR application was followed by the manufacture’s protocol (Sansure Biotech): reverse transcription at 50□ for 30 minutes, cDNA preincubation at 95_ for 1 minutes, 45 cycles of denaturation at 95°C for 15 seconds, and extending and collecting fluorescence signal at 60 °C for 30 seconds. Results definition accords to the recommendation by the National Institute for Viral Disease Control and Prevention (China) ^10^.

### Ethics approval

This study was approved by the ethics committee of the First People’s Hospital of Huaihua (KY-2020013102) and the Central Hospital of Shaoyang (KY-202000103), China. Considering the infectious of NCP, we conducted an oral informed consent with every patient instead of written informed consent (www.chictr.org.cn Chi CTR2000029734).

### Statistical analysis

Normally distributed continuous variables were described as mean and standard deviation (SD). For non-normally distributed continuous variables, we used median and interquartile range (IQR) or range. Categorical variables were expressed as ratio and percentages (%). Differences in means of normally distributed continuous variables were compared using Student’s t-test (two groups) and the non-normally distributed continuous variables compared using Mann-Whitney U test. Categorical variables were compared using the χ^2^ test or Fisher exact test. A two-sided P-value 0.05 was considered statistically significant. All statistical tests were performed using SPSS version 25.0.

## Results

### Control measures in China and current status in two centers

Since China firstly reported the outbreak of a cryptic pneumonia to WHO on December 31, 2019. The causative agent was soon identified as a novel coronavirus on Jan 7, 2020. As the sharply increased number of NCP in Wuhan, China ordered a shutdown of Wuhan city on Jan 23, 2020. Hunan Province, geographically closed to Wuhan, immediately launched a level one emergency response to prevent the infection spreading. The other cities also responded strict control measures in succession. From Jan 22, 2020 to Feb 12, 2020, a total of 104 cases were confirmed in the two centers of Hunan Province, 48 (46.15%) were imported cases and 56 (53.85%) were indigenous cases.

Since Feb 6, 2020, imported case no longer appeared in the two centers (Figure 1 A). The cumulative number of confirmed cases increased smoothly in the two centers (Figure 1 B), newly confirmed cases per day ranged from 0 to 11, a slight increase of newly confirmed cases was observed from Jan 22, 2019 to Feb 4, 2020, and then the number turned to a little decline lasted to Feb 12, 2020 (Figure 1 B). The stable of the two centers was quite different from the sharply growth of patients in Wuhan in recent month.

**Figure 1.**
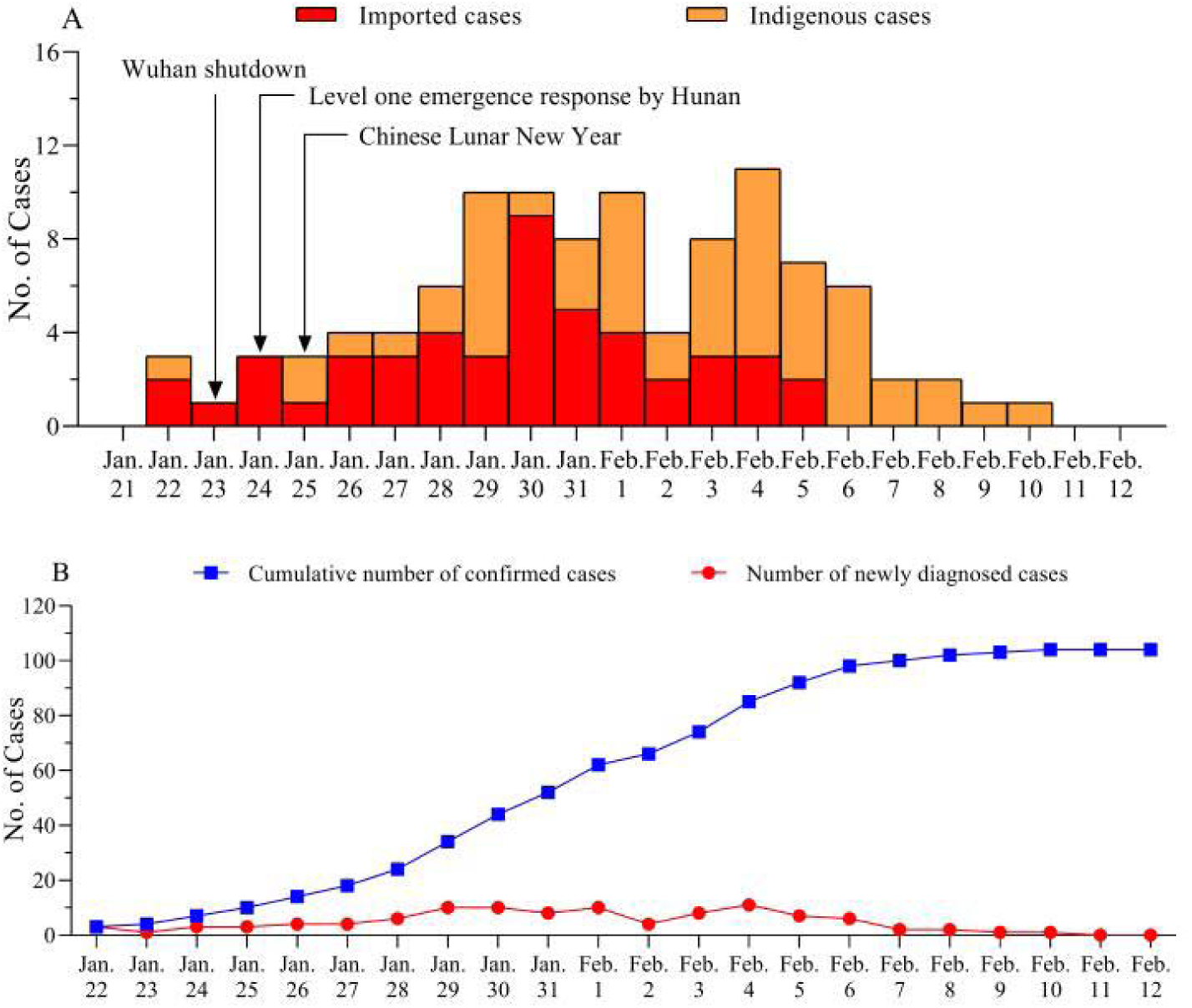
Timeline of control measures in China and current status in two centers.

**Figure 2.**
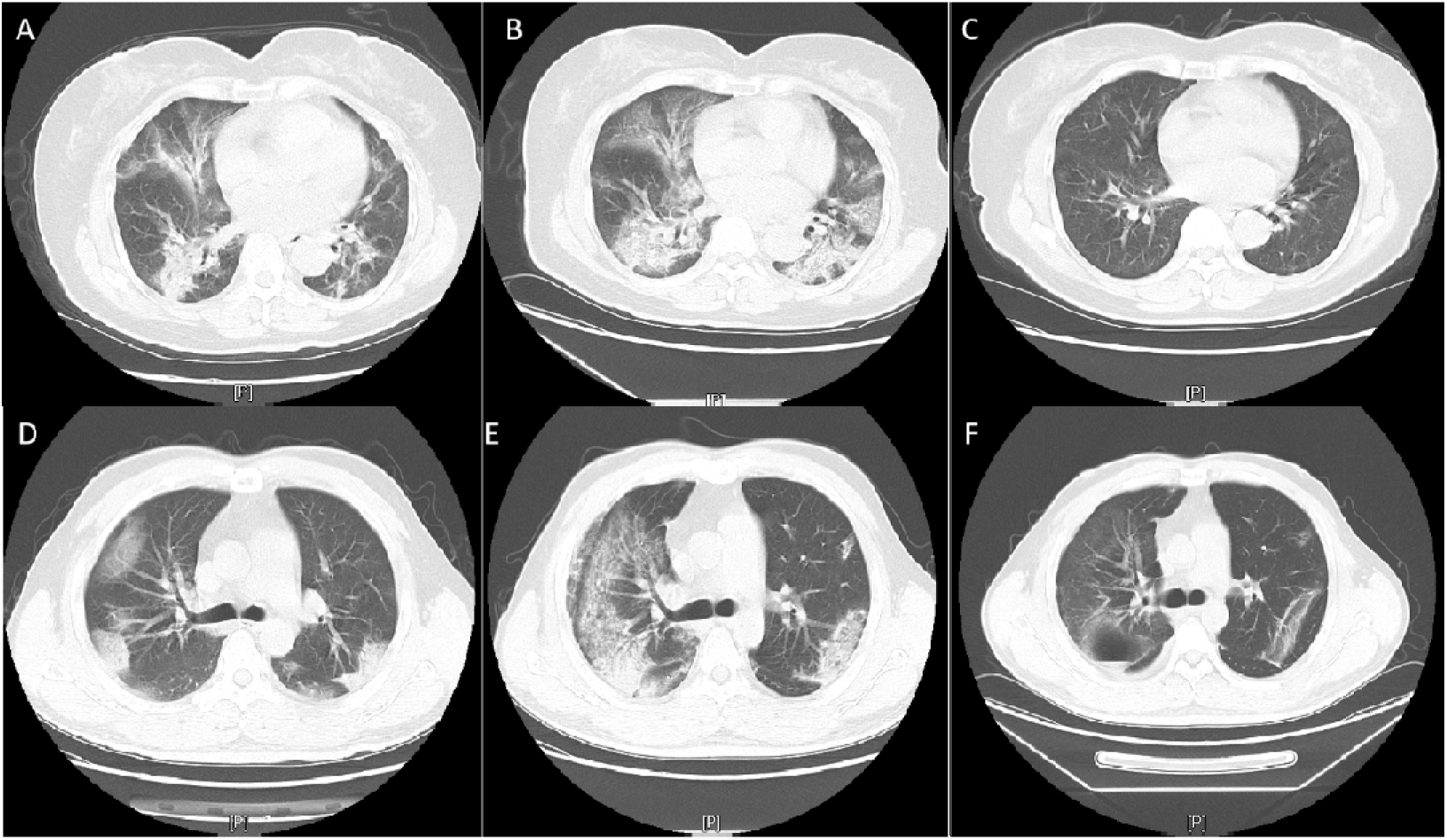
Chest Computed Tomographic Images of two Discharged Patients of Serious.

The timeline of control measures in China and the time distribution of imported cases and indigenous cases (A). The growth of cumulative confirmed patients and the newly confirmed cases per day (B).

### Transmission characteristics of 104 patients

With the aim to better understand the transmission characteristics of COVID-19 in outside-Wuhan cities. We carefully clarified the contact history of each patients. Of the 104 patients, 93 (89.42%) patients had a clear contact history with the infections, 11 (10.53%) were sporadic cases that hardly identified a definite contact history. As showed in Table 1, cluster infections including couples, relatives, friends and colleagues transmitted through a close domestic life or dinner. Family clusters accounted the most infections of COVID-19 in this study population. Cluster 6 (2 cases) and 14 (7 cases) infected via taking the same public vehicle together. Nosocomial transmission did not happen so far in the two centers.

**Table 1.**
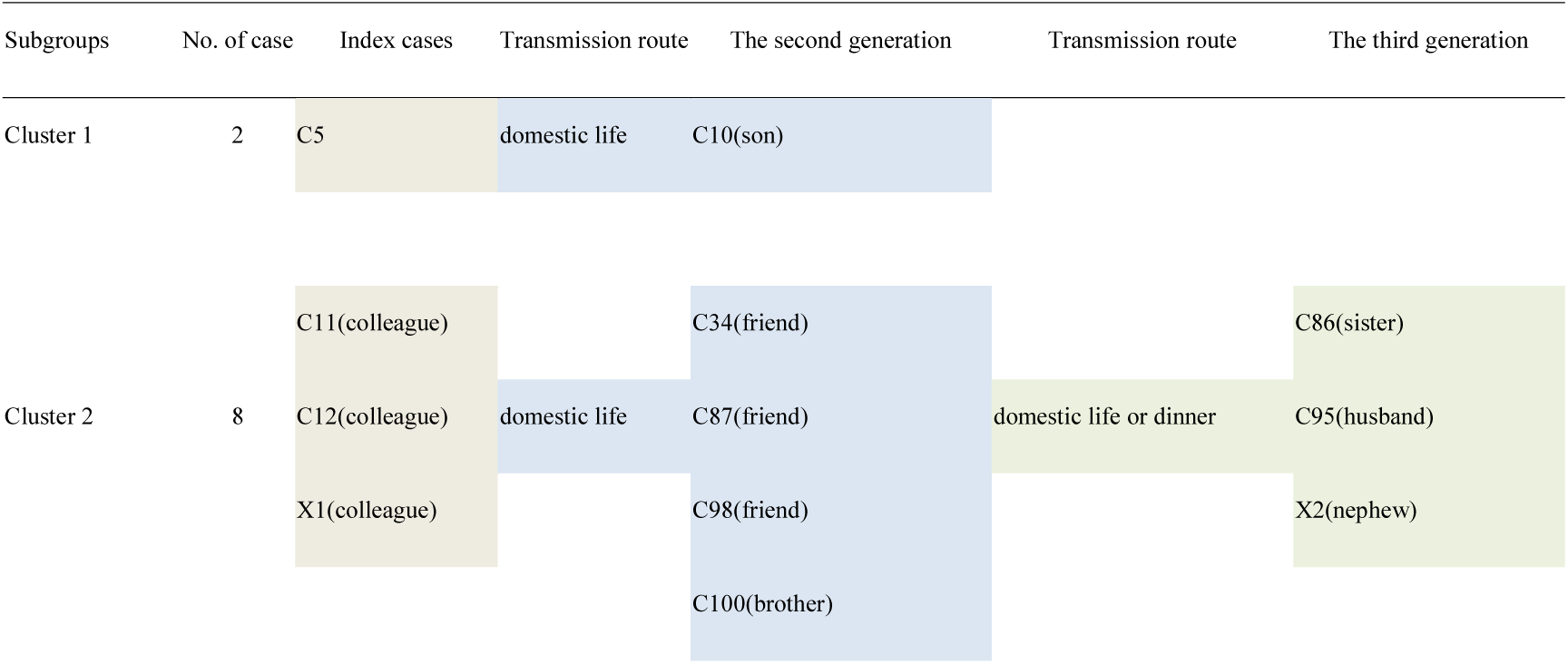
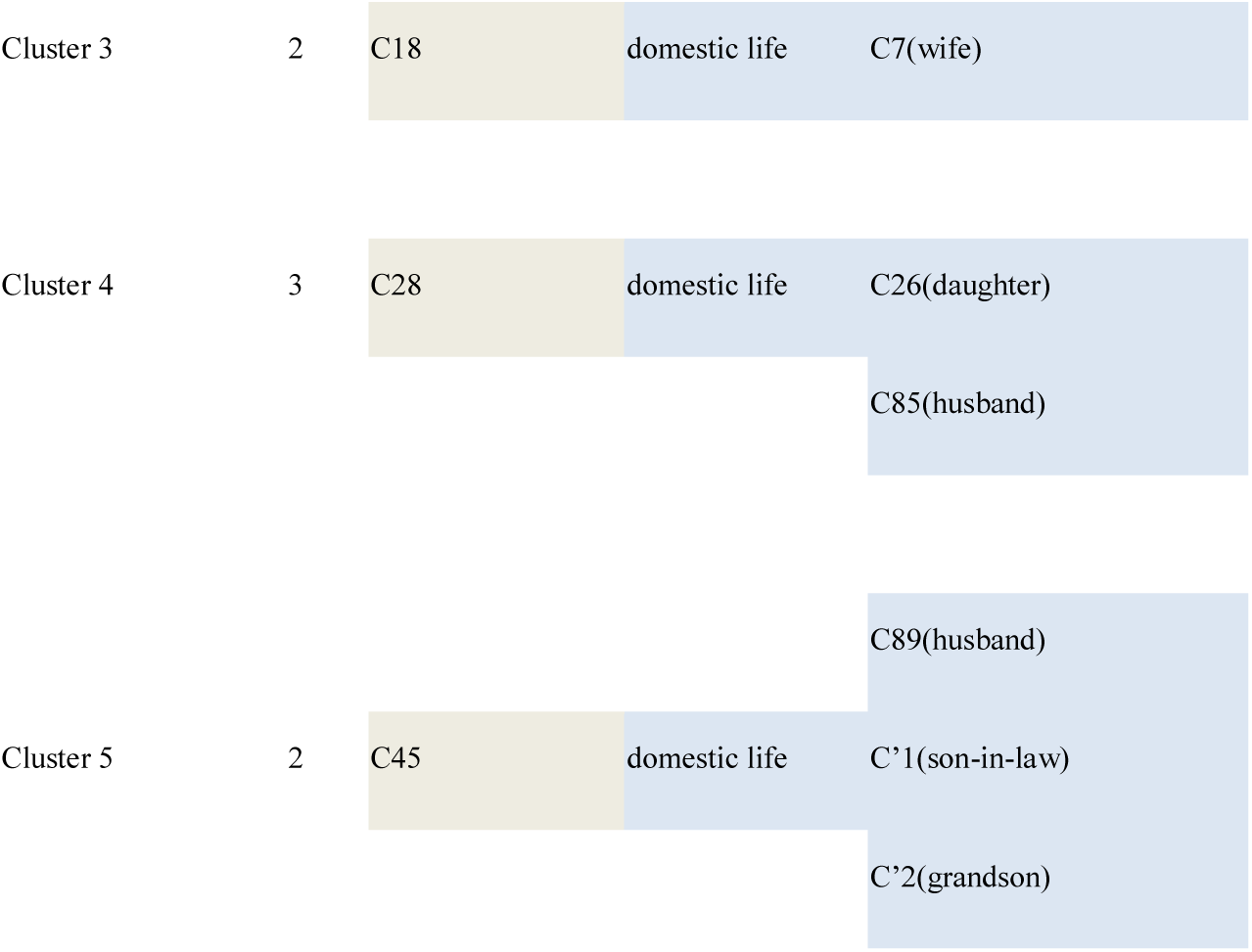
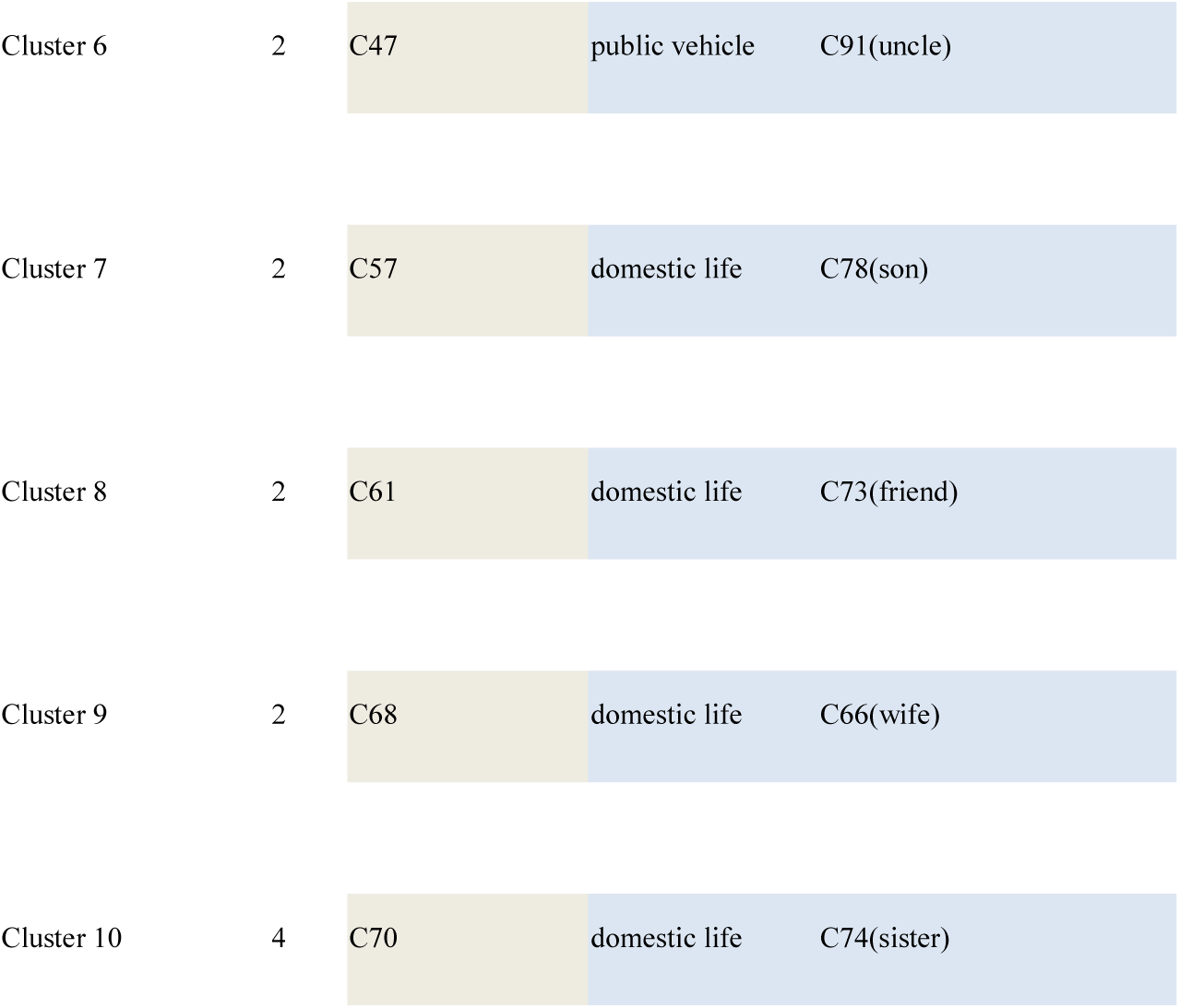
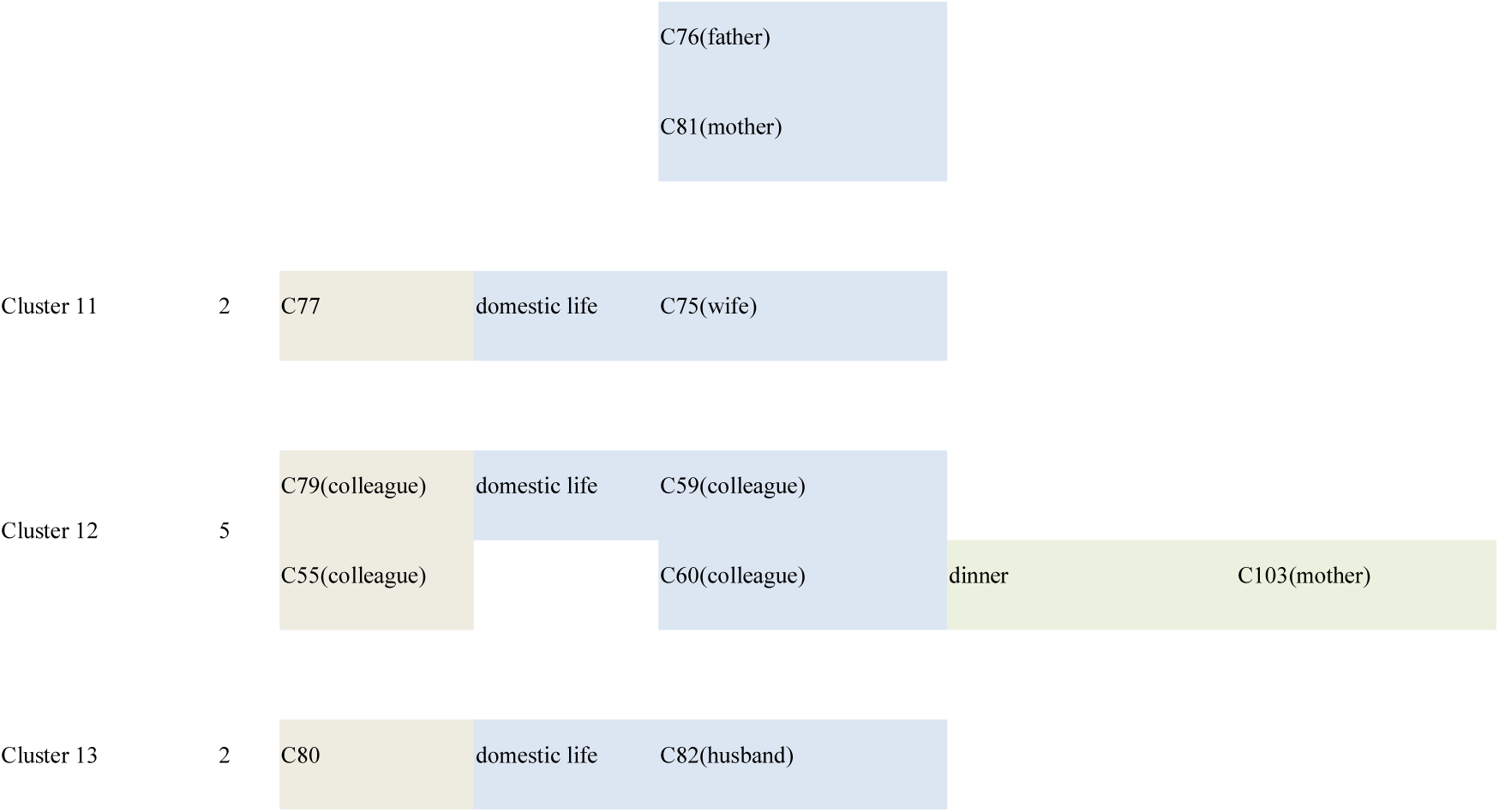
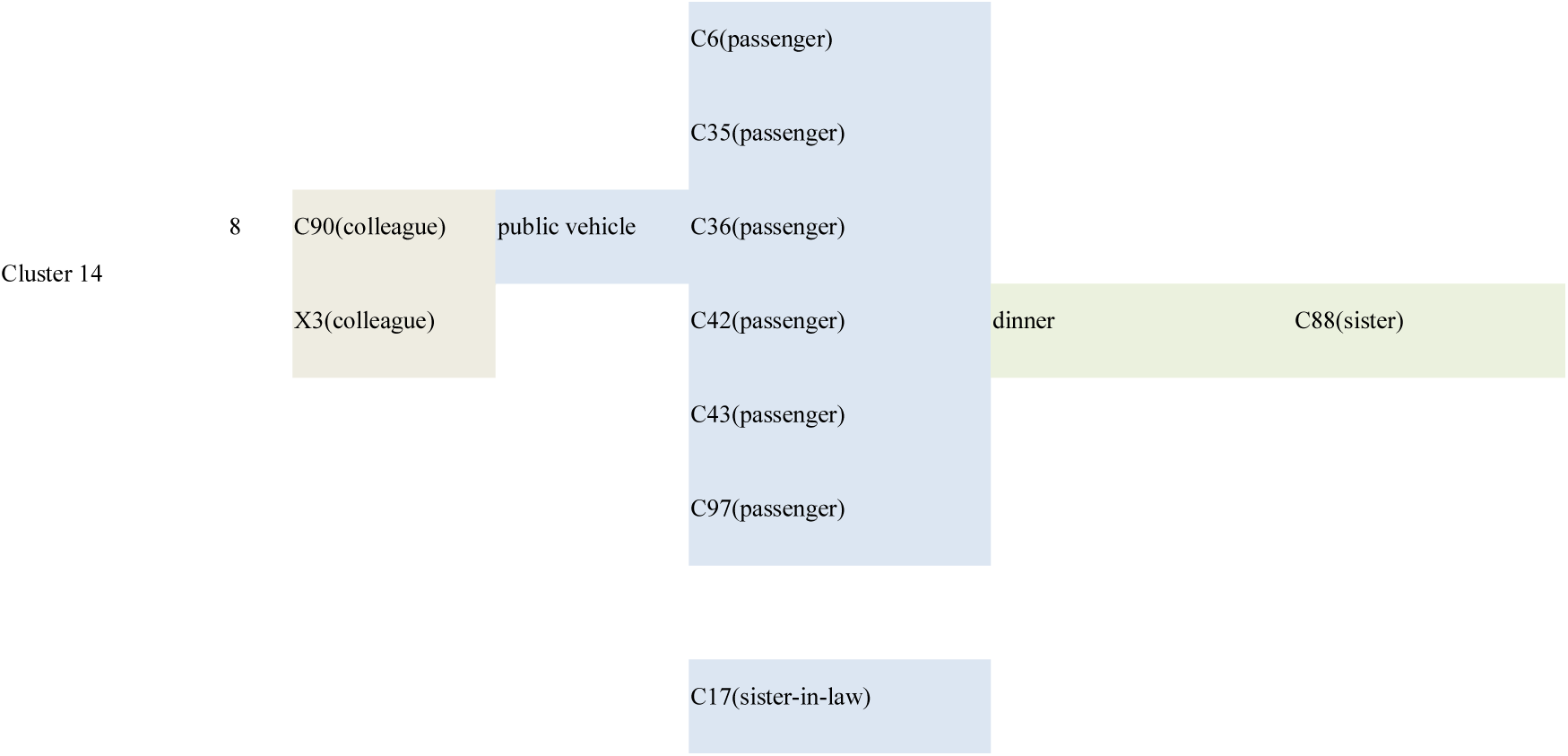
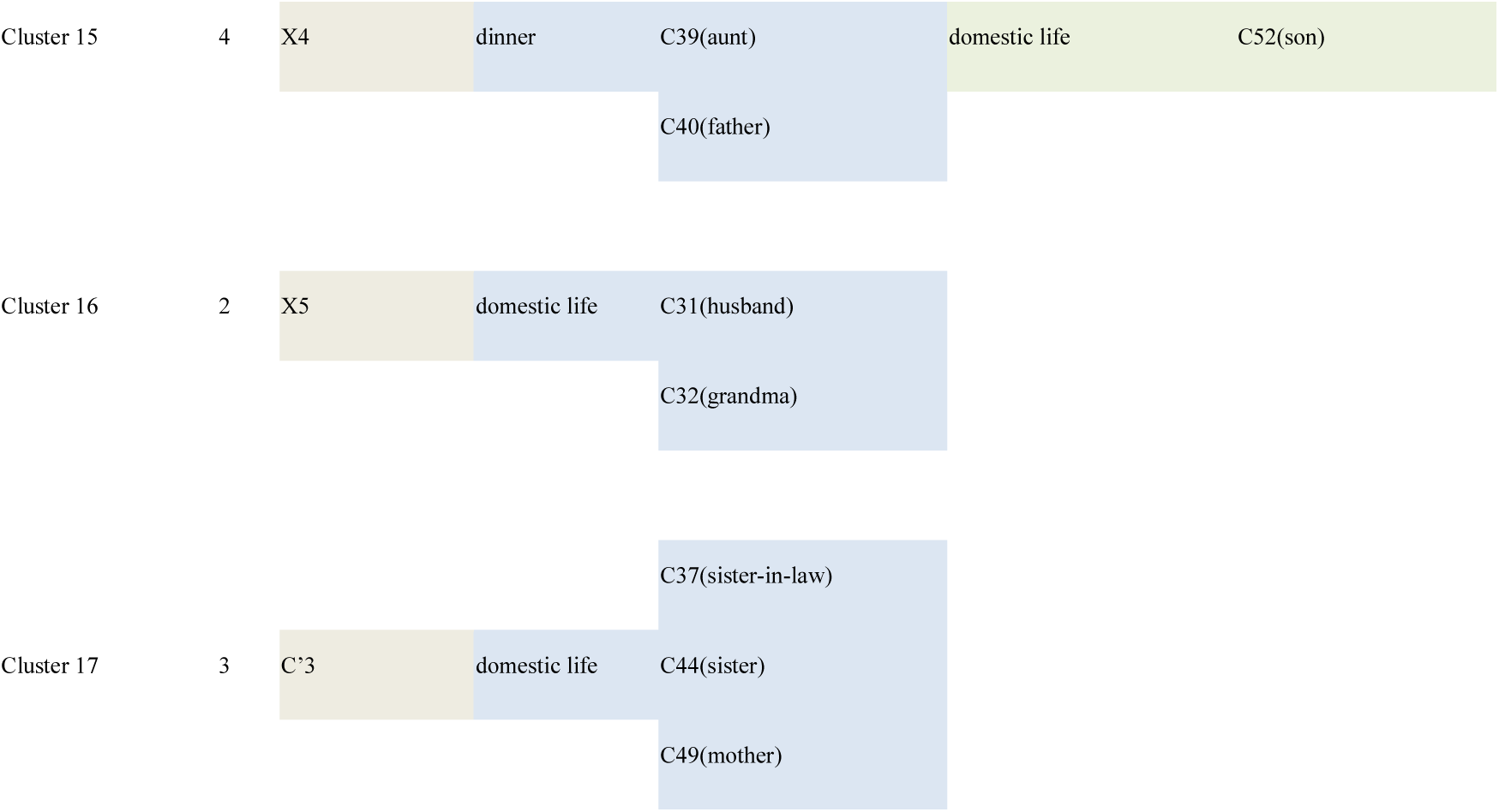
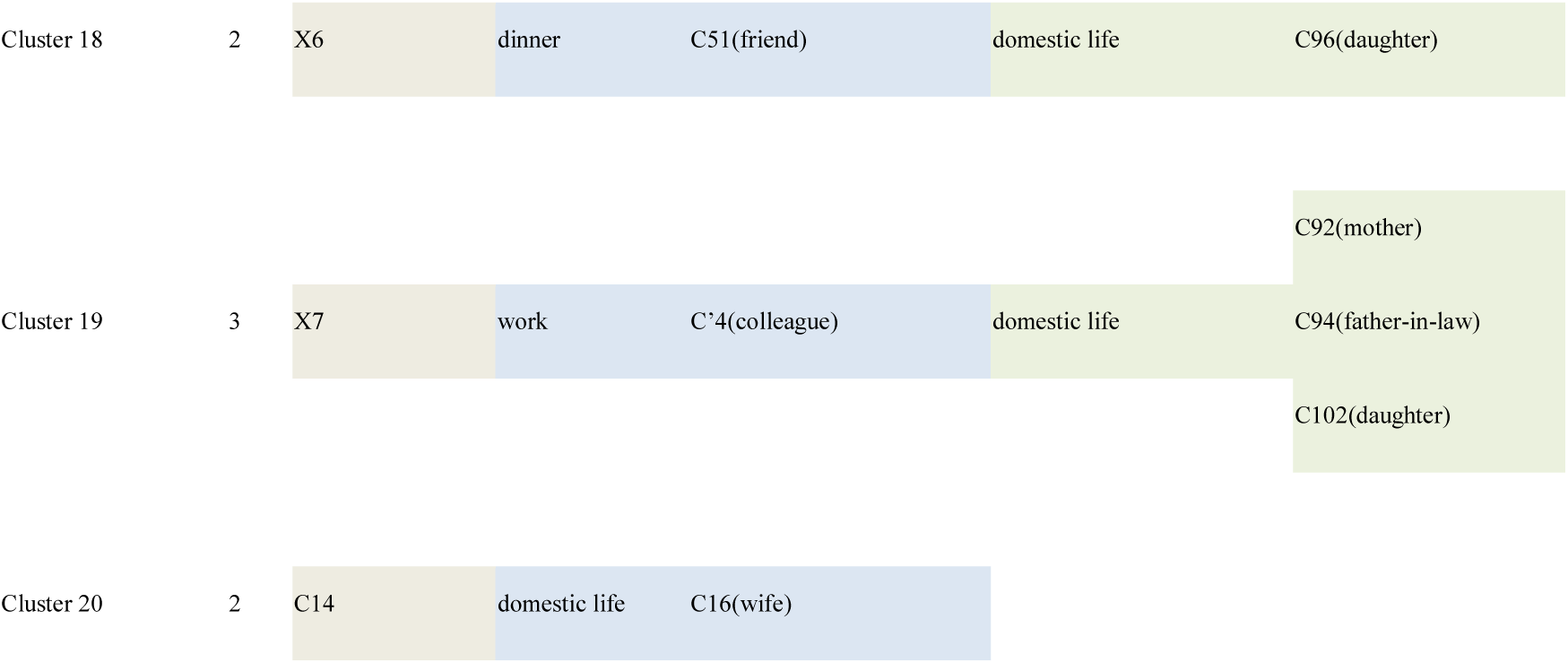
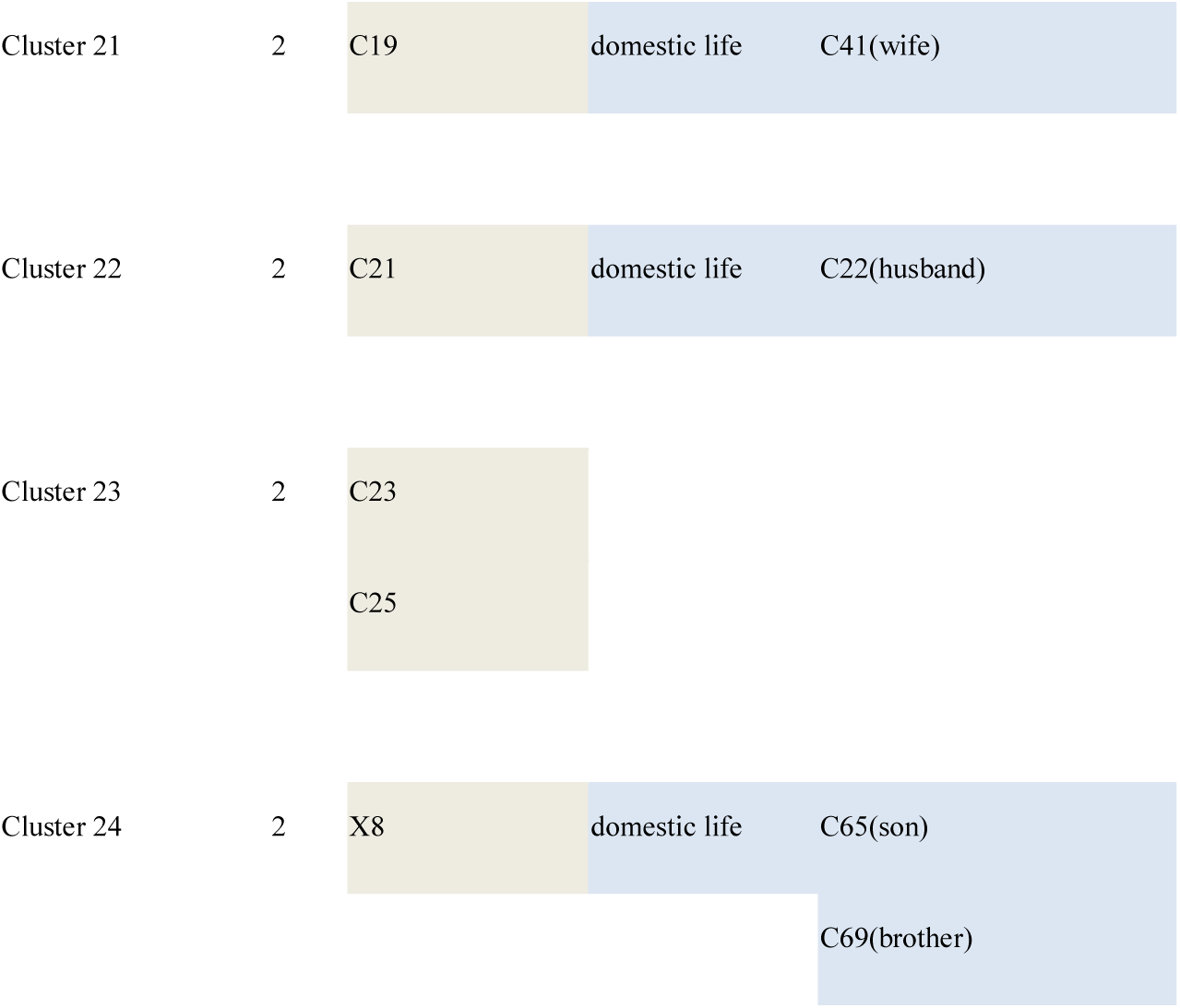
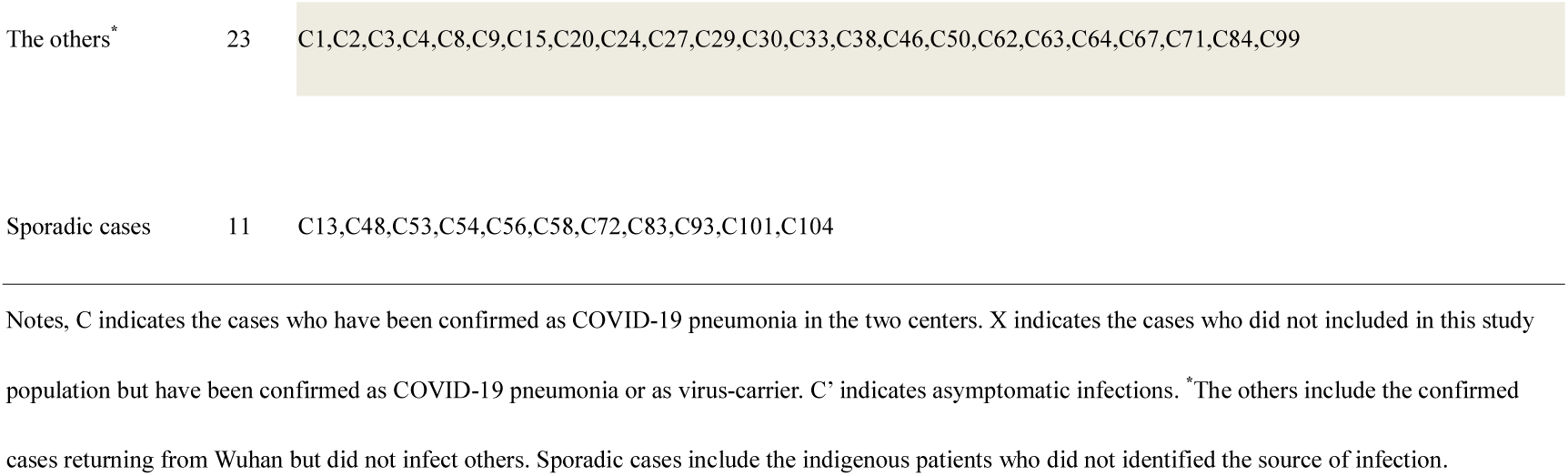
Transmission characteristics of 104 patients.

Six clusters (Table 1, cluster 2, 12, 14, 15, 18 and 19) demonstrated the existing of transmission chain of 3 “generation” (index case of one cluster identified as an infector who originally contracted the COVID-19 from Wuhan and then infected someone else, who infected another individual). Of note, 5 asymptomatic cases (C’1, C’2, C’3, C’4 and C102) were found in this study. In cluster 5, C89 was infected from his wife C45. With the aim to fast screen the potential infections, their family members took the PCR test. Their son-in-law (C’1) and their grandson (C’2) (C’1 and C’2 not included in this study population) got positive results in another hospital, but till now all of them had never developed any symptoms. In cluster 17, C’3 (not included in this study population) returned Shaoyang city from Wuhan on Jan19, 2020, three relatives of C’3 were identified as COVID-19 infection after several days of closely contacted with C’3. None of them had contacted with the other suspected infectors during those days. Her sister-in-low (C37) was confirmed on Feb 1, 2020, her sister (C44) and mother (C49) were confirmed on Feb 4, 2020. But so far C’3 had never developed any symptoms. Weather C’3 is an asymptomatic infection did not been identified by PCR test, but the same contact history and the similar onset time of her three relatives indicate that C’3 was an asymptomatic COVID-19-carrier. In cluster 19, C’4 (not included in this study population) contacted with her college who traveled from Wuhan, and soon confirmed by PCR positive result. As an asymptomatic patient, C’4 infected C92 (C’4’s mother), C94 (C’4’ s father-in-law) and C102 (C’4’ s daughter), C102 also had no symptoms with a positive result of PCR test.

### Clinical characteristics and medical treatment of 104 cases

As showed in Table 2, of the 104 patients, 49 (47.12%) were male, the mean age was 43 (SD, 7.54, rang, 8-83) years, three children with NCP aged 13, 8 and 8 years. There were 22 (21.15%) patients that had one or more comorbidities on admission, diabetes (12 [11.54%]), hypertension (15 [14.42%]) and cardiovascular disease (7 [6.73%]) were the most common comorbidities. Common onset symptoms included dry cough (79[75.96%]), fever (66[63.46%]), expectoration (39[37.50%]), fatigue (33[31.73%]), muscular soreness (20[19.23%]) and dyspnea (15[14.42%]). A few patients developed diarrhea (2[1.92%]) and palpitation (1[0.96%]) as onset symptoms. The median incubation duration was 6 days, ranged from 1 to 32 days; 8 patients got more longer incubation duration (18, 19, 20, 21, 23, 24, 24 and 32 days) that more than 14 days. Median time from onset to confirmation was 6 (rang, 0-17) days. There were 16 (15.38%) patients were identified as severe, the ratio of male vs. female was 11:5 and median age was 53 (rang, 18-81); 9 (8.65%) patients required ICU care, the ratio of male vs. female was 4:5 and median age was 59 (rang, 18-84). Of the 9 ICU patients, 3 received invasive ventilation and 4 received noninvasive ventilations. Some patients presented with organ function damage, including 5 (4.81%) with liver function abnormal, 3 (2.14%) developed with cardiac injury and 2 (1.89%) developed with acute kidney injury. ARDS occurred in 13 (12.50%) patients. For the period from admission to developed ARDS, the median time was 2 (1-8) days. As we followed until Feb 12, 2020, 40 (38.46%) had discharged and 1 (0.96%) died, the rest 63 (60.58%) patients stayed in hospital.

**Table2.**
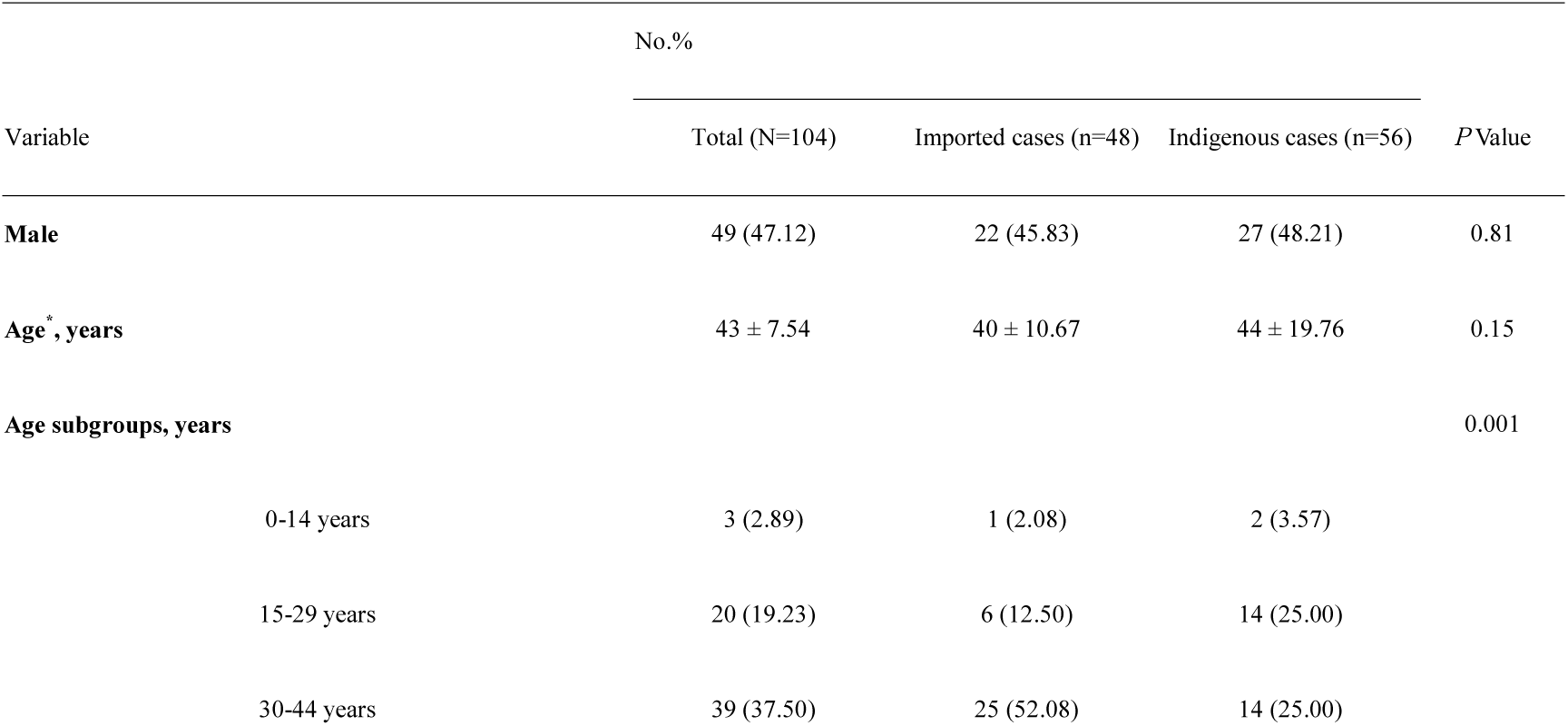
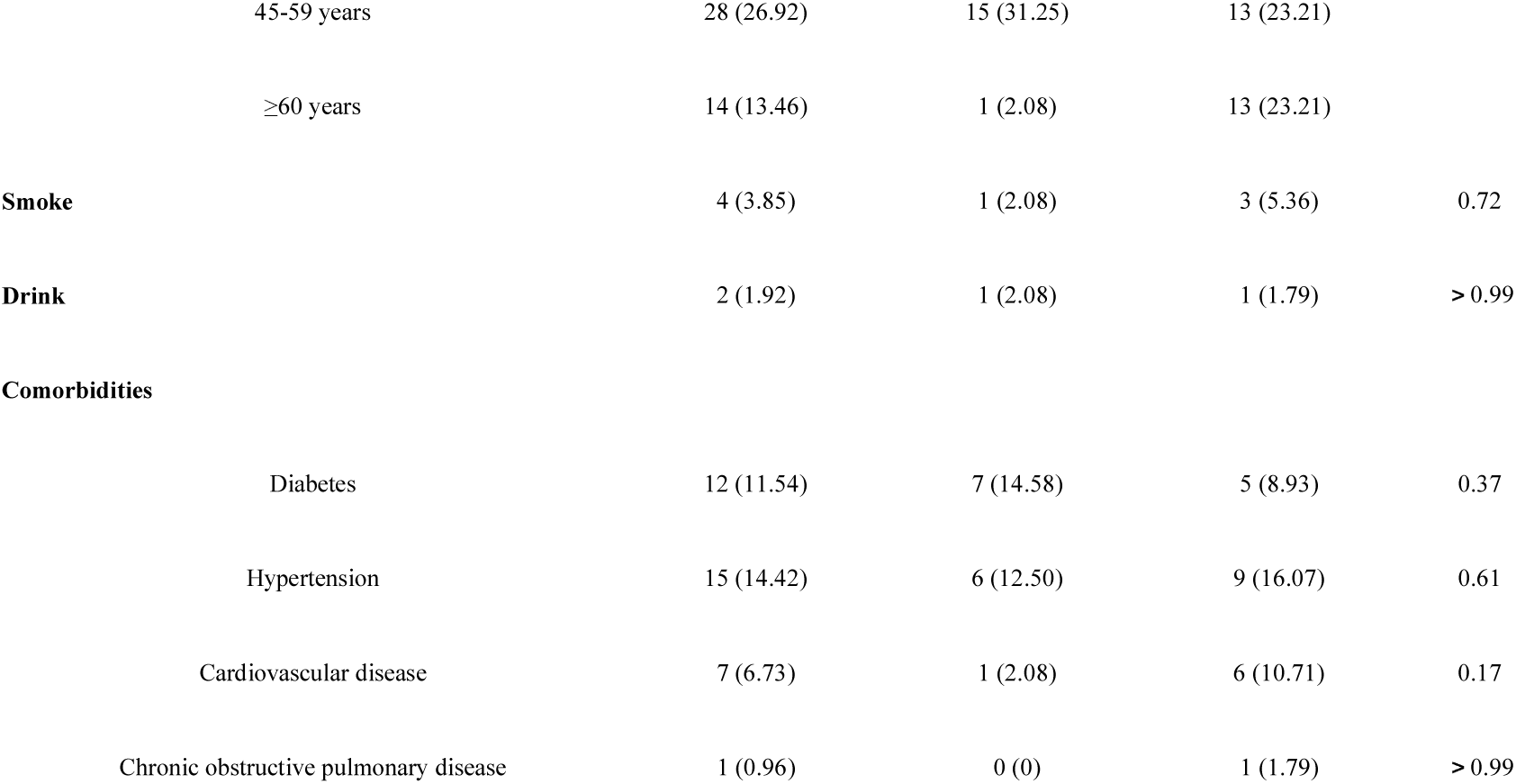
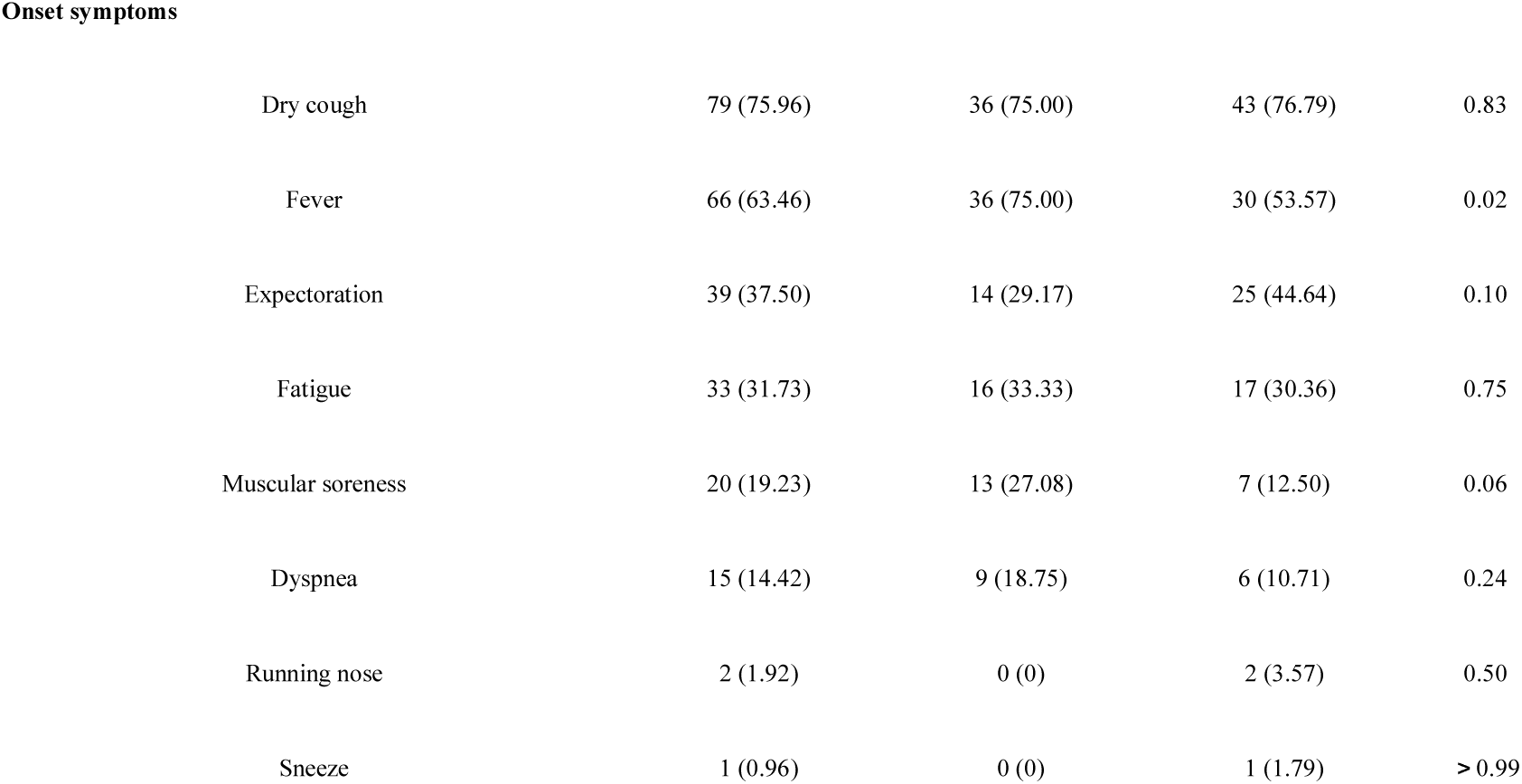
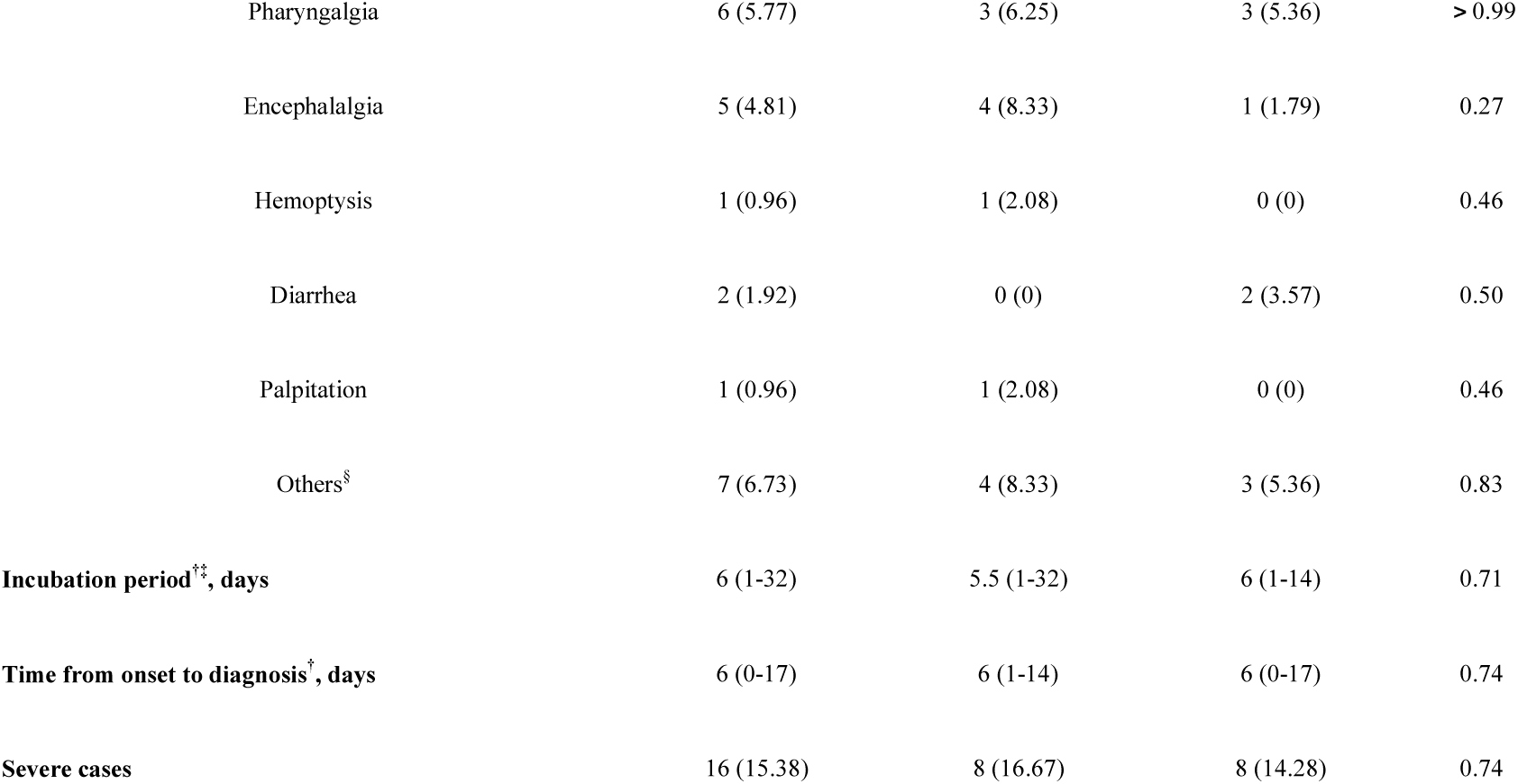
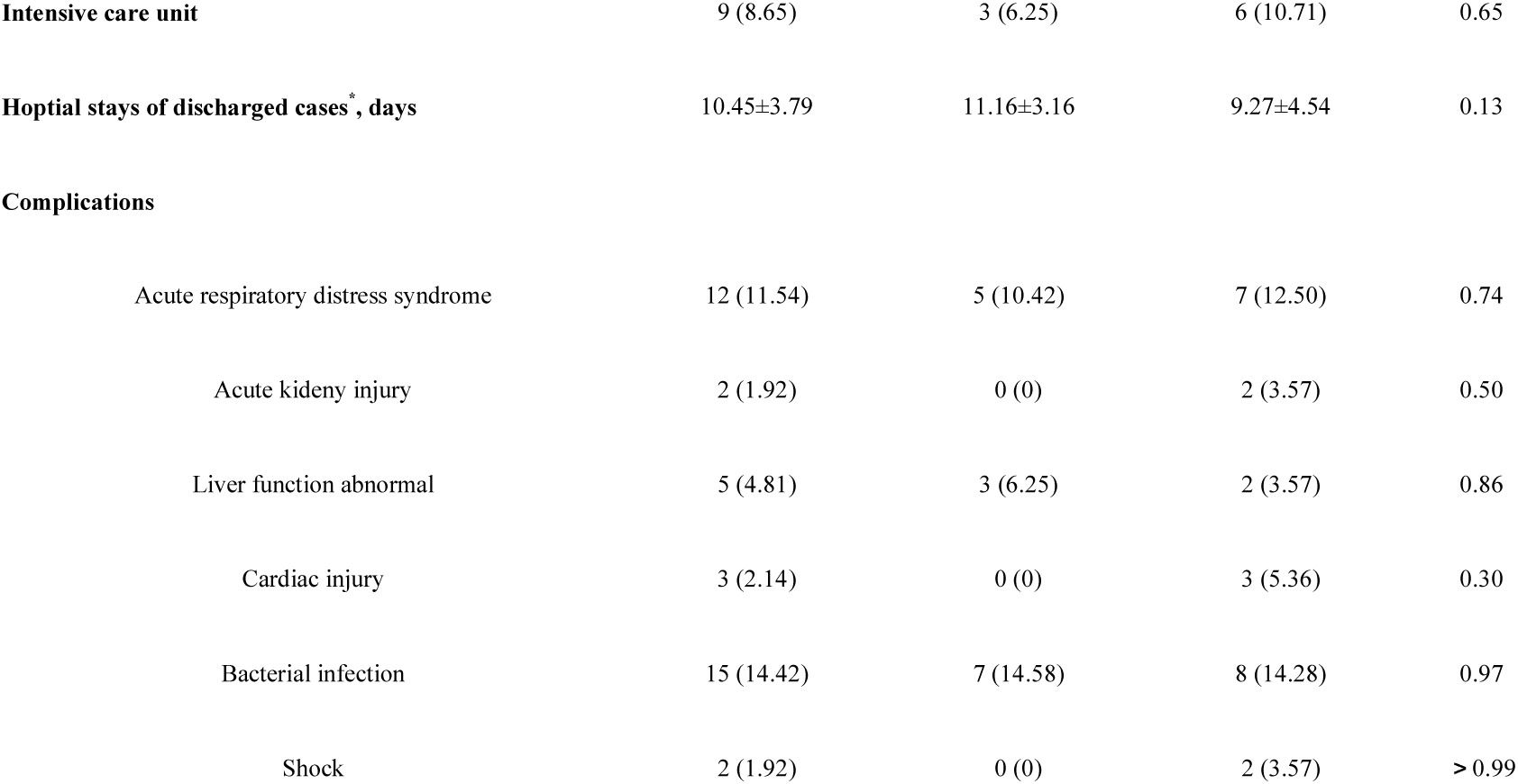
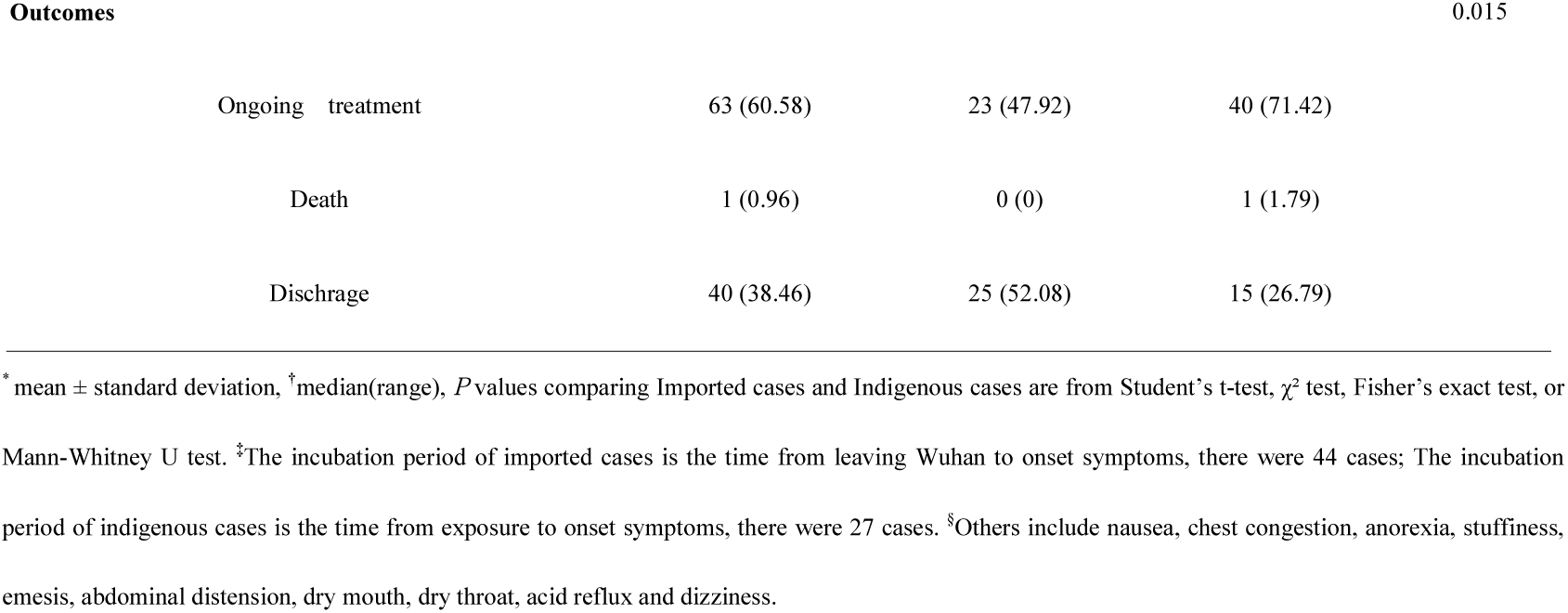
Demographics and clinical characteristics of patients infected with COVID-19

As showed in Table 3, for the white cell count, 71 (70.30%) of 101 patients was normal, 15 (14.85%) were below the normal and 15 (14.85%) were above the normal level; 101 patients took the lymphocyte count test, results showed 37 (36.63%) patients were normal and 63 (62.38%) patients were below the normal level; 82 patients took the CRP test, 29 (35.37%) were normal and 53 (64.63%) were above the normal level; 101 patients took the PCT test and all were at the normal level; 70 patients took the test of erythrocyte sedimentation rate (ESR),results showed 17 (27.14%) patients were normal and 51 (72.86%) patients were above the normal level; 25 patients took the IL-6 test, 15 (60%) patients were normal and 10 (40%) patients were above the normal level. For the D-dimer test, 56 (56%) of 100 patients were normal and 44 (44%) were above the normal level. Just 19 patients took the CD4 and CD8 count test, 17 patients had lower level of CD4 and 9 patients had lower level of CD8. Compared with the imported patients, indigenous patients had higher lymphocyte counts, lower level of triglyceride, total protein and globulin, no significant differences of the other laboratory parameters between the two group was observed. 103 (99.04%) patients received antiviral therapy, 51 (49.04%) received 1 to 2 antiviral drugs, 52 (50.96%) received 2 or more antiviral drugs (Interferon α atomization 97 [93.27%], Lopinavar/ ritonavir 86 [82.69%], Abidol 66 [63.46%]). Some (21[20.19%]) received antibacterial therapy, glucocorticoid therapy (9 [8.65%]), immunopotentiation therapy (thymalfasin (9 [8.65%]) and immunoglobulin (14[13.46%]). It was worth pointing out that 80 (76.92%) patients received traditional Chinese medicine therapy.

**Table3.**
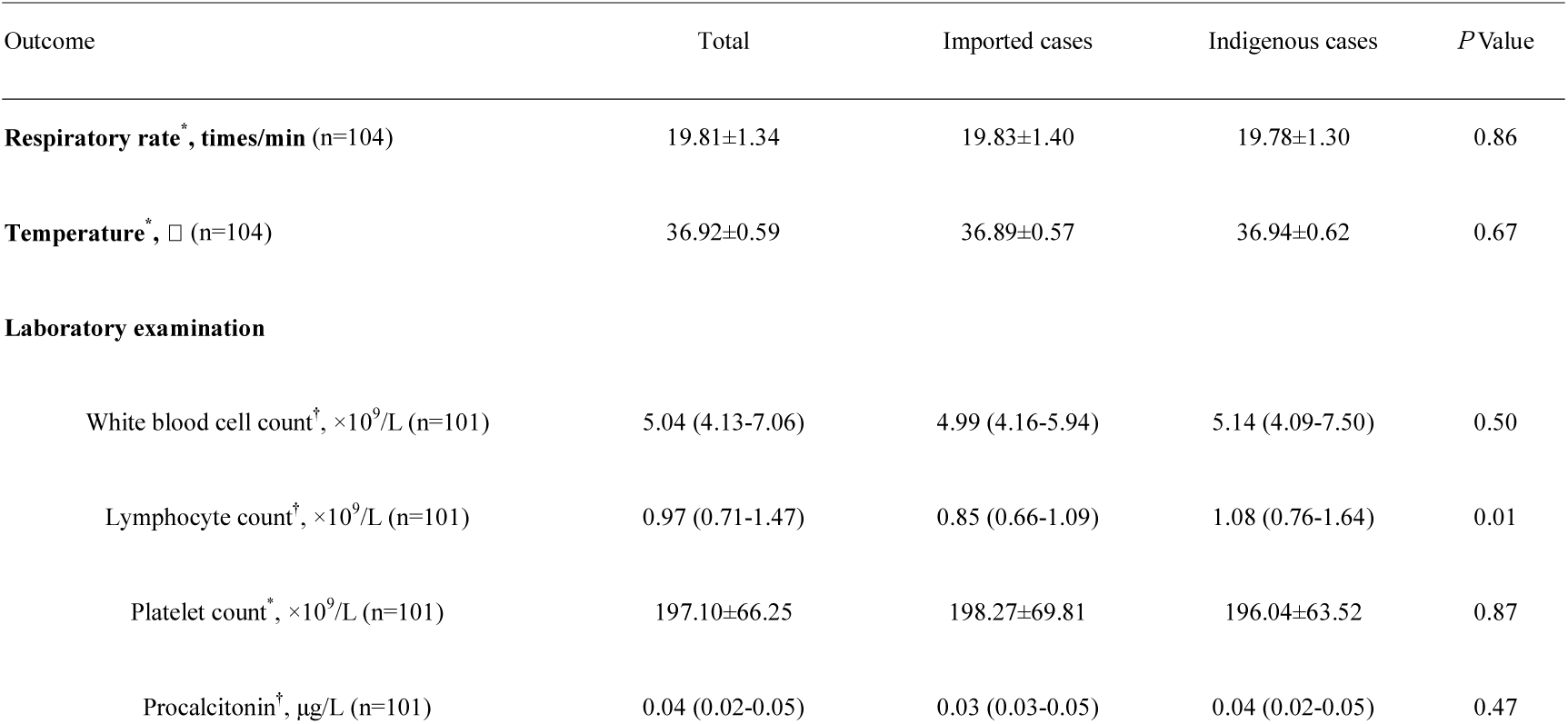
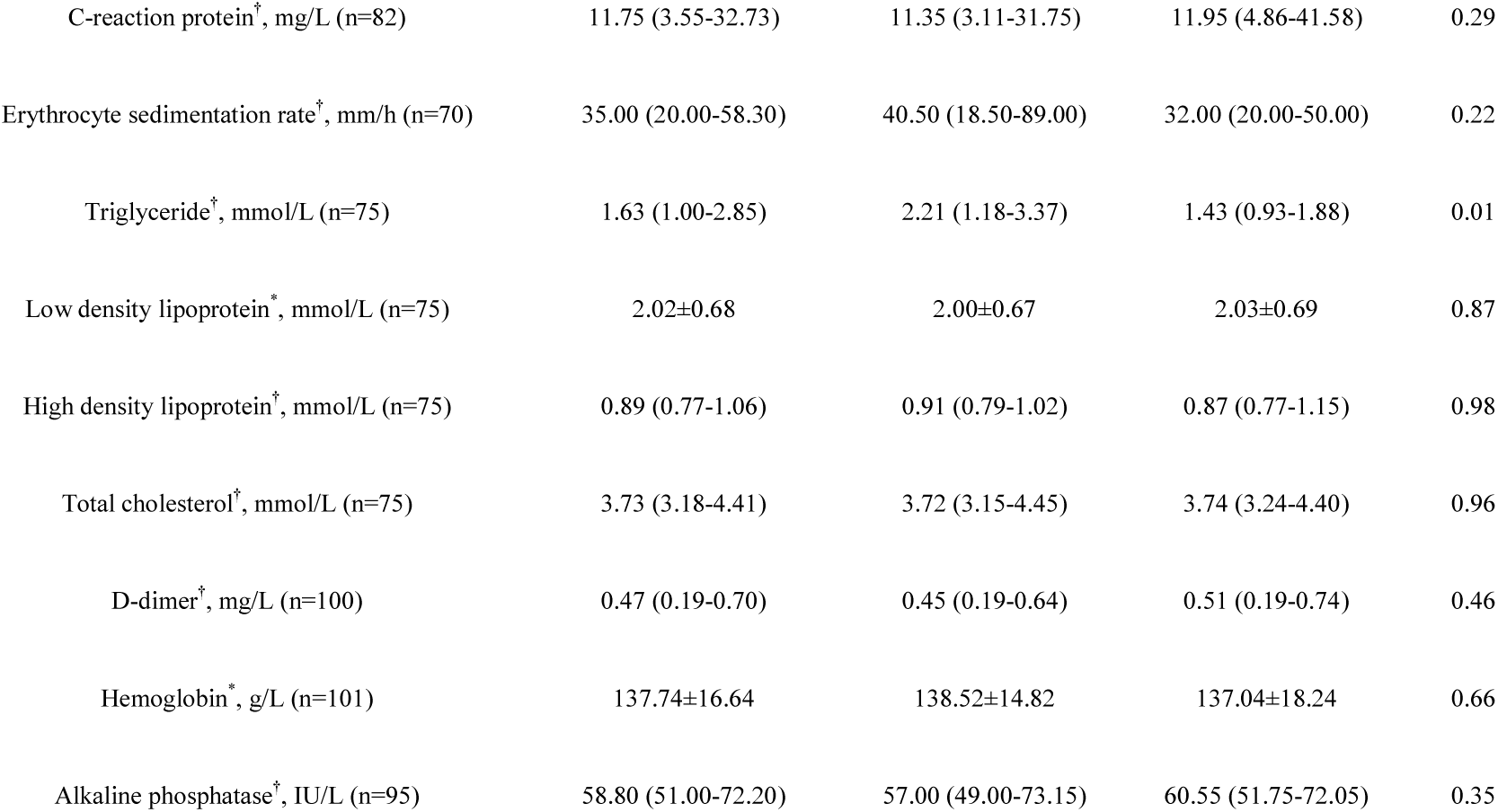
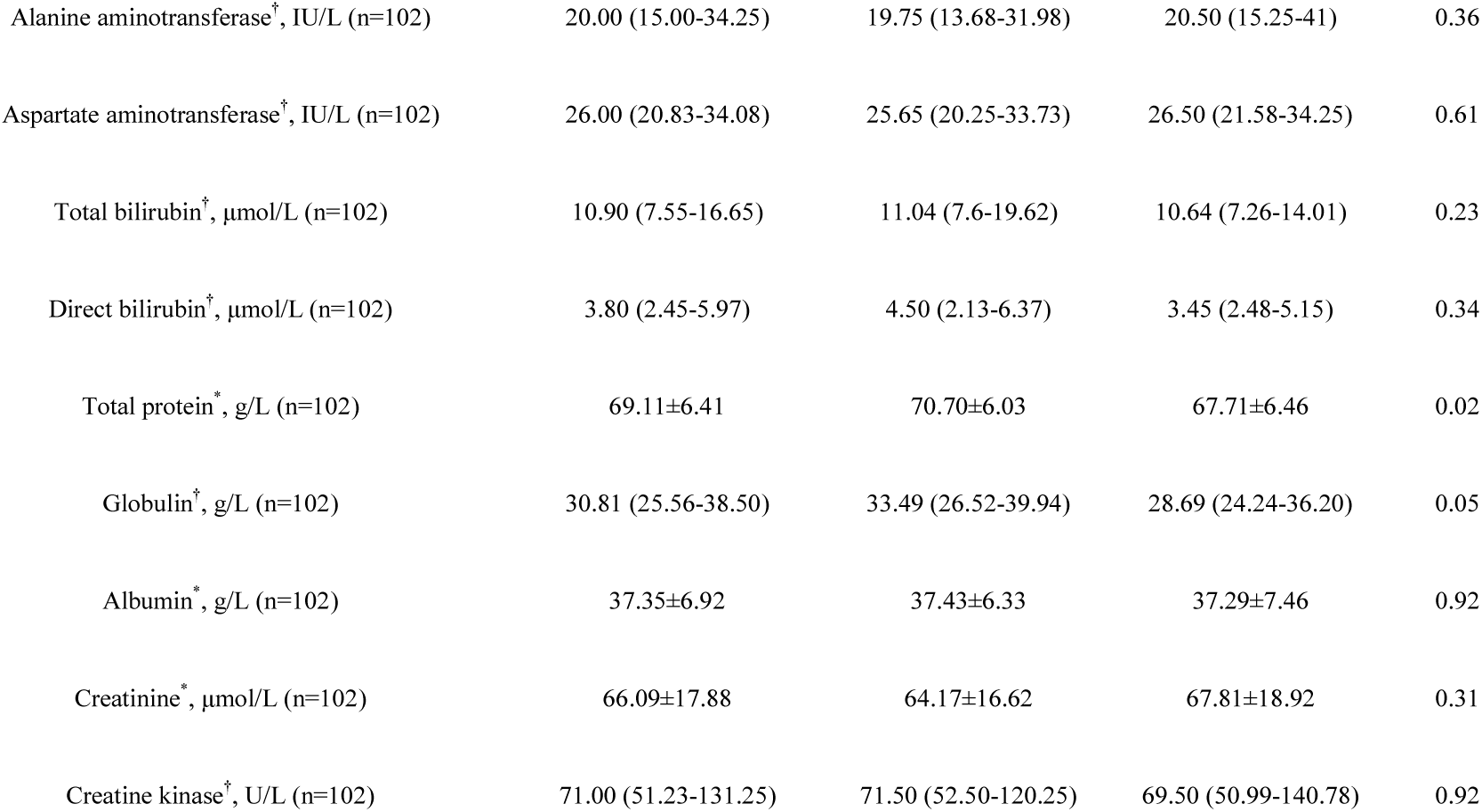
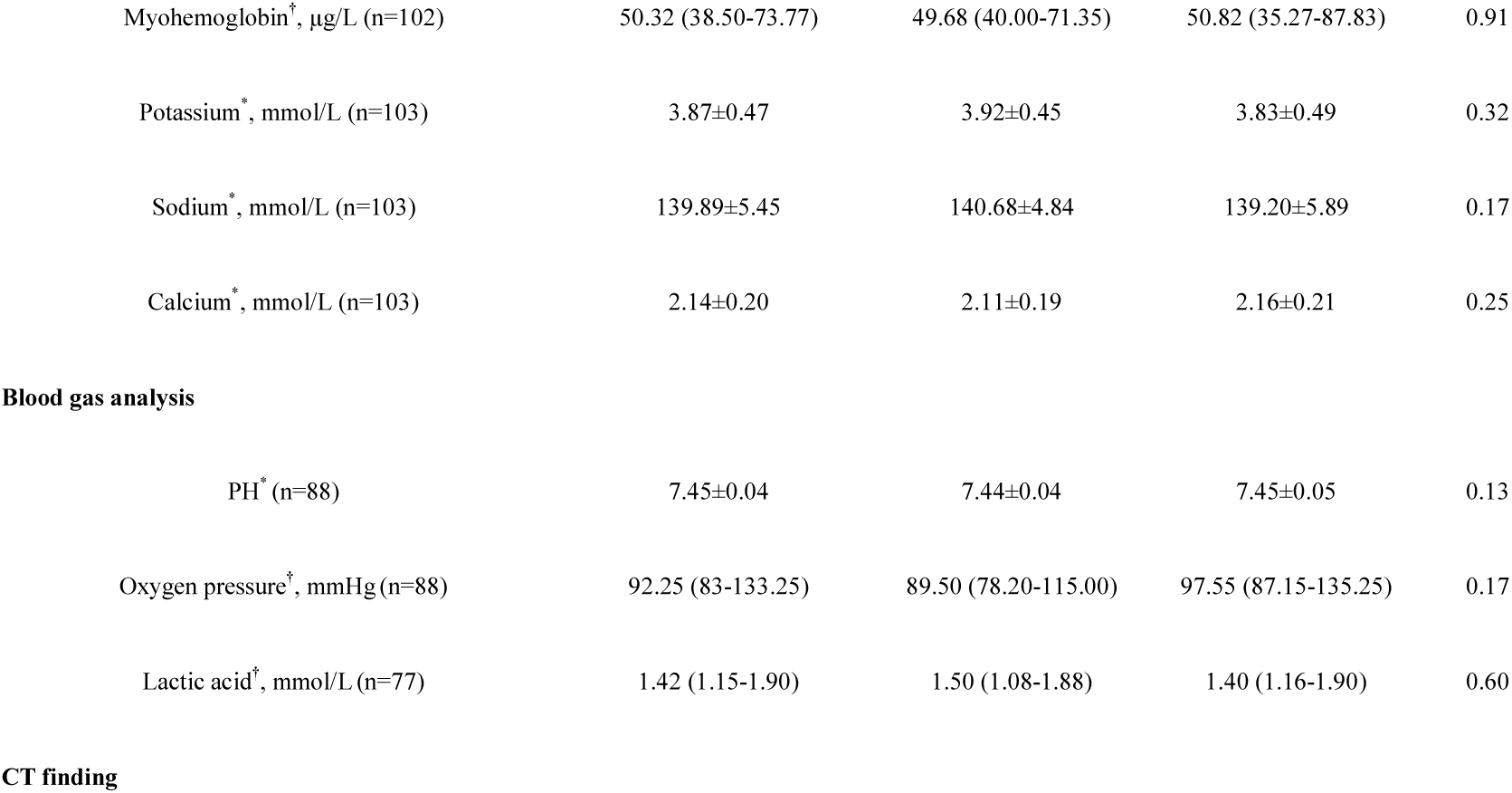
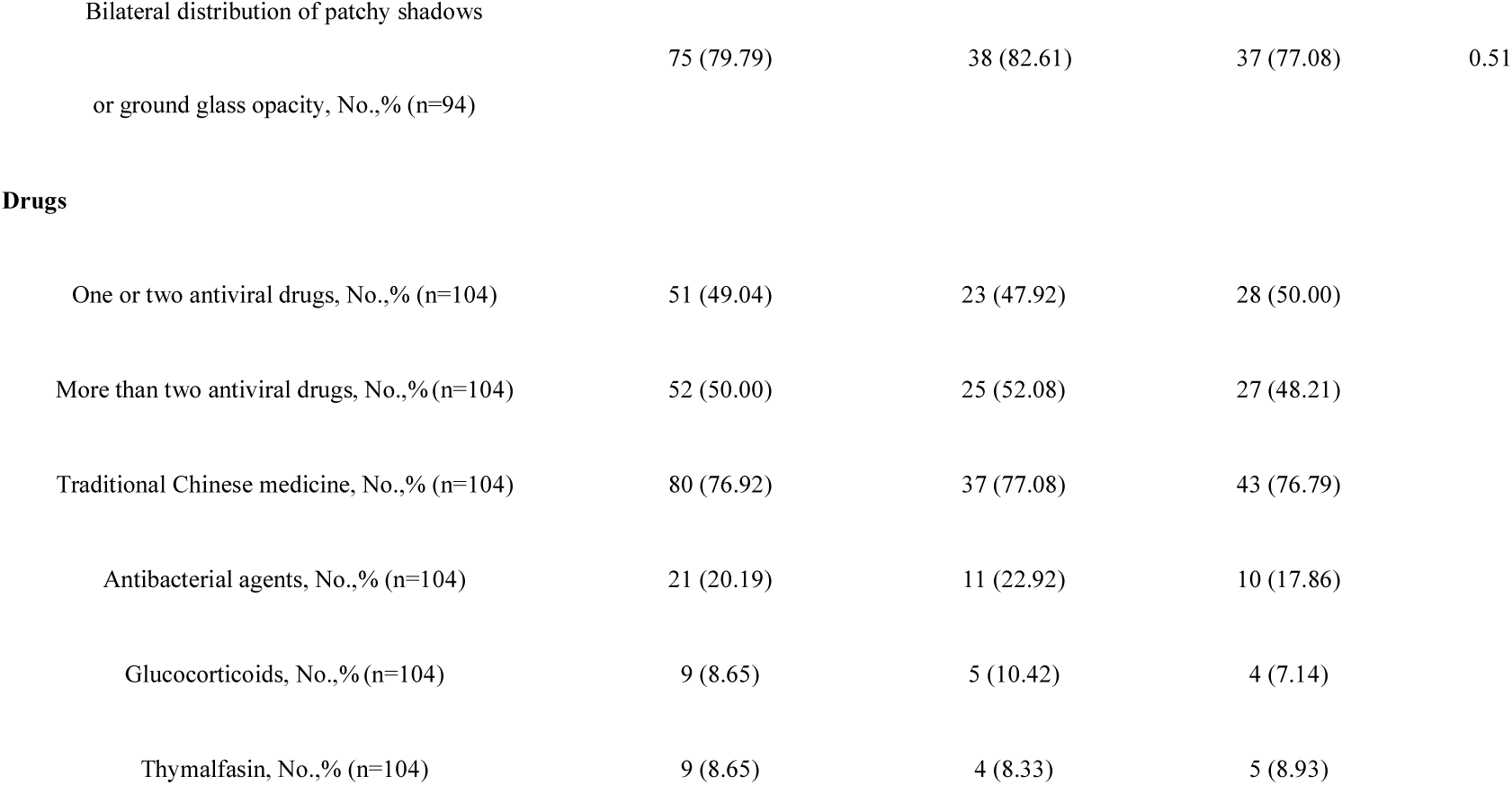
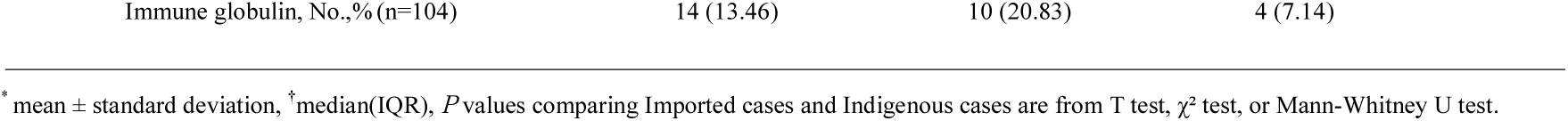
Laboratory results (patients on admission to hopital) and CT findings of patients infected with COVID-19

### CT imaging findings of two discharged patients of serious conditions

There were 75 of 94 (79.79%) patients lesions involving both lungs. Data of initial chest thin-section CT imaging findings in two discharged patients (C56, Figure 4 A, B, and C; C54, Figure 4 D, E and F) with NCP were showed here. CT images on admission (Figure 4 A, D) showed multiple patchy pure ground glass opacity (GGO), GGO with reticular and/or interlobular septal thickening. After the disease progressed at 7 and 9 day (Figure 4 B, E), it was found multiple ground glass shadows and infiltrating shadows in bilateral lung involvement, even more consolidation lung lesions occurred. In addition, lung parenchyma from peripheral to central lung interstitium, no findings included pleural effusion. On the day 21 of admission (Figure 4 C, F), most of the lesions in the lungs were absorbed and some of them showed changes of pulmonary interstitial fibrosis.

### Conditions Infected With NCP

A-F. Baseline CT images at admission of the first discharged patient (IP: C56, 57 years) show multiple patchy pure ground glass opacity (GGO), GGO with reticular and/or interlobular septal thickening (A). After the disease progressed, Follow-up CT images at 7 day show pulmonary consolidation occurred, lung parenchyma from peripheral to the central lung interstitium (B). Follow-up CT images at 21 day show most of the lesions were absorbed and some of them showed changes of pulmonary interstitial fibrosis (C).Baseline CT images at admission of the second discharged patient (IP: C54, 37years) show multiple patchy pure ground glass opacity (GGO), GGO with reticular and/or interlobular septal thickening (D). After the disease progressed, Follow-up CT images at 9 day show pulmonary consolidation occurred, from peripheral pulmonary parenchyma to the central lung interstitium (E). Follow-up CT images at 21 day show most of the lesions were absorbed and some of them showed changes of pulmonary interstitial fibrosis (F).

## Discussion

Here we report the transmission and clinical characteristic of COVID-19 pneumonia in 104 outside-Wuhan patients. The main observations of this case series were followed. First, the smoothly increase in the cumulative number of confirmations of the two centers indicates that the timely control measures work well. Second, family clusters represent as the major body of infections, transmission along the chain of 3 “generations” was observed. The asymptomatic transmission exists and may bring more risk to the spread of COVID-19. Third, no gender difference of patients was found, indicating male and female may have the same susceptibility of this illness. Fourth, clinical characteristics of this study population were similar to the previous reported patients.

Since the outbreak in Wuhan, it is unclear how many people truly infected and how many infected people of Wuhan has imported to the other cities. Screening the potential infectors related to Wuhan was implemented quickly by the local governments after the announced of Wuhan shutdown. Dynamic analysis of 104 cases from two centers in Hunan province showed that the imported cases mainly appeared during Jan 22, 2019 to Feb 5, 2020, then the indigenous cases became the major body, suggesting the initial cases have been founded over nearly 20 days of screening. Our observation showed a smoothly increase in the cumulative number of confirmations, newly confirmed cases per day also remained relatively stable. Compared with the contemporary growth trend in Wuhan, explosive increase in the confirmed cases was not observed in the two centers from Jan 22, 2019 to Feb 12, 2020, indicating that the timely control measures in the two cities worked well. But the true turning point of decline remained to be observed. While the massive travel of returning after Spring Festival season, continued strict control measures is very important to block the infection spreading.

Yet fundamental information gaps exist on how to accurately assess the transmission efficiency. While the controversy of sharply increased cases and medical shortage in the early and outbreak stage in Wuhan, patients in Wuhan may have limitation to fully reflect the true epidemiological characteristics of this illness. Evidence has suggested person-to-person transmission of COVID-19 via droplets or skin touch^2,3,11^. The data of this study showed a notable feature is clustering occurrence, most patients were infected from their family members, relatives or friends through a close contact. Only 11 (10.53%) of this study patients were sporadic cases that hardly identified infector source, suggesting that community transmission of COVID-19 is not developed rapidly in the two cities (Huaihua and Shaoyang); this also matches the smooth growth of total confirmed cases. Of note, strict control measures by the local government produced a powerful effect on the slowing spread.

We are eager to know how infectious the virus is. Except the confirmed cases, whether the asymptomatic COVID-19-carriers has the infectious is unclear. Three cases (C37, C44 and C49) infected from the same person (C’3) who ever traveled to Wuhan. But until now, C’3 did not develop any symptoms. Though we did not take a PCR test to confirm whether C’3 was a virus-carrier, the same contact history and the similar onset time of her three relatives indicate that C’3 was an asymptomatic COVID-19-carrier. Five asymptomatic patients were found in this study, one patients (C’4) who infected three family members (C92, C94 and C102) provide evidence that the asymptomatic transmission risks the spread of COVID-19, which brings more difficult to cut off the epidemic’s transmission route. We surveyed eight infected couples, a total of 3 infants were closely lived with their parents, but none of them was infected. Just 3 children were infected from their parents or relatives. These observations further demonstrated that infant and child are not so susceptible as adult, that is consistent with the previous reports^2,3,6,12^. Unlike the other reported populations, no nosocomial transmission was found in the two centers^2^. The safeguard of protective equipment and the strengthen of nosocomial infection control may play key roles in the zero accident of hospital-related infection.

Unlike some earlier reports^3,6^, here no gender difference was found among this study patients (47.12% patients were male). This is consistent with a recent report of 138 Wuhan patients ^2^. This report further provides the evidence that male and female may have the same susceptibility of this illness. This study patients were younger than that of reported patients. It may be related to the patients’ job characteristics and social relationship. With the Spring Festival coming, young or middle-aged people are more likely to attend social activity, which results in person-to-person transmission. Common symptoms of onset were similar to the reported patietns^1-3^. The atypical symptoms such as diarrhea, nausea and runny nose bring us more difficult to diagnose precisely. The incubation duration ranged from 1 to 32 days with the median time of 6 days which was similar to the reported patients^13^. A recent report warned us the incubation duration may extend to 24 days^14^. We also found the incubation duration of 8 patients ranged from 18 to 32 days, indicating that it may exceed 14 days which reported with the initial infections ^3^.

Patients who required ICU care just presented 8.65%. With the increased awareness of early discovery and timely treatment, organ function damage was occurred just in few patients, that is quite different from observation of patients in Wuhan patients. The higher rate of discharge (38.46%) and lower mortality (0.96%) of this study population may mainly attribute to the relatively superior treat conditions, including enough healthcare worker and single ward for every patient. In addition, psychological intervention was also performed to patients. Studies suggested that COVID-19 may attacks human’s immune system which resulted in cytokine storm^3^. The lymphocyte counts of this study patients were below the normal. Here 17 of 19 patients showed a significant decrease in CD4 cell counts and 9 of 17 patients showed a decrease in CD8 cell counts, it is a pity that rest of the patients did not take the test. We still don’t know the pathogenic mechanism of COVID-19, so we should take a route test of the CD4 and CD8 counts for better understanding of this illness.

Though no antiviral treatment for COVID-19 infection has been proves to be effective^15,16^. Antiviral and supportive treatment are the major therapy for NCP. 103 of 104 patients of this study received one or more antiviral drugs, including Lopinavar/ ritonavir, interferon α atomization and Abidol. Lopinavar/ ritonavir was proved to be substantial clinical benefit against SARS^17^ and MERS^18^. Particularly, 80(76.92%)patients received traditional Chinese medicine therapy, the efficacy in COVID-19 treatment needs more clinical evidence to be confirmed. Controversy about corticosteroids in NCP treatment has not reached a consensus^19,20^. Evidence suggests corticosteroids did not decrease the mortality of patients with SARS and MERS, but rather delayed the clearance of viral^19,21^. Chinese guideline recommends a short treatment of corticosteroids in server NCP^7^. Just 8.65% patients of this study received glucocorticoids treatment, most of them are severe patients.

Baseline CT images of the cases showed most pulmonary lesions involved bilateral lungs with multiple lung lobes, with predominant distribution in posterior and peripheral part of the lungs^20^. No difference was found from previous studies^2,3,6,22^. Lung consolidation may occur in severe cases, therefore, finding of consolidation lesions would serve as an alert in the management of patients.

This study has several limitations. First, just two centers of Hunan Province were included, there is limited information based on the data to fully assess the transmission and clinical characteristics in outside-Wuhan cities. Second, all patients were confirmed by RT-PCR through nasopharyngeal or throat swab, it could not reflect viral load change in blood or organs. Until now it is confused about whether the severity of NCP is related to changes of viral load in blood. Third, the follow-up period is not long enough to examine the outcomes of all the included patients.

In conclusion, this report gives showed that timely control measures after the Wuhan shutdown blunted the spreading of COVID-19 in other cities. Family but not community transmission occupied the main body of infections in the two centers. Asymptomatic transmission demonstrated here warned us that it may bring more risk to the spread of COVID-19. The incubation period of 8 patients exceeded 14 days.

## Data Availability

Data will be obtained from China clinical trial registry(Chi CTR2000029734)after six months of article publishing

http://www.chictr.org.cn

